# Public understanding, awareness, and response to monkeypox virus outbreak: A cross-sectional survey of the most affected communities in the United Kingdom during the 2022 public health emergency

**DOI:** 10.1101/2022.08.25.22279207

**Authors:** Sara Paparini, Ryan Whitacre, Melanie Smuk, John Thornhill, Chikondi Mwendera, Sophie Strachan, Will Nutland, Chloe Orkin

## Abstract

**Objectives:** To examine awareness, understanding, and response to the monkeypox (MPXV) outbreak among the most affected communities in the United Kingdom.

**Methods:** A cross-sectional survey was administered between June 15 and July 27, 2022, targeting adults using LGBTQI community channels on social media platforms and the dating app, Grindr. Basic descriptive statistics have been created and logistic regression implemented to explore associations. Odds ratio p values have been given for completeness. No formal power calculation was implemented.

**Results:** 1932 respondents completed the survey, 1691 (90%) were cis or trans men who have sex with men. 1750/1932 identified as men, 88 as women and 64 as gender non-conforming. 1453 (77%) described their sexual identity as gay/ lesbian, 64 (3%) as queer, 221 as bisexual (12%), 81 (4%) as heterosexual, 37 (2%) as pansexual. 744 (39%) were under the age of 40. 71% (1366) self-identified as being from White, 3% (67) Black, 8% Asian (158), 2% LatinX(36) and 11% (203) ‘Mixed or Other’ heritage groups. 85% were employed and 79% had completed higher education. 137 (7%) identified themselves as living with HIV when asked if they lived with a long-term condition. Overall, 34% reported a limited understanding of public health information, 52% considered themselves at risk, 61% agreed people infected with MPXV should isolate for 21 days, 49% reported they would first attend a sexual health clinic if infected, 86% reported they would accept a vaccine and 59% believed MPXV originated from animals. Reported level of understanding of public health information, agreement to the need to isolate for 21 days, vaccine acceptance, and beliefs about the origins of MPXV varied by income level, education and heritage. Trust in healthcare providers, perception of risk of MPXV, and willingness to be vaccinated differed by gender and sexual identity. The most trusted sources of information were healthcare professionals (37%), official health agencies (29%), and mainstream media (12%). Younger participants were most likely to state they would not engage with clinical services if they had MPXV symptoms.

**Conclusions:** In this group of respondents, largely made up of White, male, employed men who have sex with men with university education, vaccine acceptability was very high, yet public health information and advice were neither universally accepted nor correctly understood. Social determinants of health inequalities already shaping the UK public health landscape are at risk of being compounded in this new emergency. Engagement with structurally disadvantaged members of affected communities is essential for the public health response. Further research is needed to establish awareness, risk perception, engagement in care, and vaccine acceptability, especially in more diverse and vulnerable populations.

## INTRODUCTION

Mounting an effective response to the human monkeypox virus (MPXV) public health emergency requires targeted public health programmes that address the needs of the most affected communities. As we have witnessed in related viral pandemics, including HIV and COVID-19 respectively, the proliferation of misinformation and processes of marginalisation pose significant barriers to care(1), contributing to delays in diagnosis, treatment, and vaccine uptake, and thus compounding health inequalities.(2-5) Yet, people who have been made vulnerable due to structural processes of marginalisation continue to be blamed for viral spread.(6)

The US Centers for Disease Control and Prevention (CDC), World Health Organisation (WHO), and UK Health and Security Agency (UKHSA) have adopted a public health approach centring around community engagement of communities of gay, bisexual and other men who have sex with men (GBMSM), demonstrating a clear understanding of the lessons learned from previous pandemics.(7) This approach is well-supported by research on the co-production of knowledge and resources with affected communities, which can deepen and facilitate engagement and support the response.(8) Indeed, leveraging knowledge gained from communities can strengthen ‘structural competency’, help counter processes of marginalisation, and contribute to strengthening systems of care and service provision.(9-11) For these reasons, public health leadership has emphasized that creating trust is essential for sustaining interventions in the response to the current MPXV emergency.

The UK national government and health authorities are playing a key role in the ongoing response. Since the first UK cases of MPXV infection were reported, the national government and National Health Services have published information online (UK.gov, NHS.gov), covering key facts about transmission, risk factors, self-isolation measures, where to attend for diagnosis and information on vaccines. This information specifies sexually active GBMSM are at highest risk for MPXV.

However, limited research is available on the success of public health interventions in the current MPXV emergency, and little is currently known about how this public health information has been interpreted, especially by GBMSM as the most affected community. A survey of a nationally representative panel of U.S. adults (n = 1,580) found nearly 1 in 5 adults (19%) were worried about MPXV infection, and the majority (60%) had limited knowledge of vaccine availability. It also revealed concerning findings about misinformation and conspiracy theories. (12). A study on MPXV information on Twitter found high levels of misinformation gaining traction (13). An online survey designed to assess understanding and acceptance of public health information in MSM (who were predominantly users if HIV pre-exposure prophylaxis, or PrEP) in the Netherlands showed similarities in intention to self-isolate and take up a vaccine amongst at-risk groups (PrEP and non-PrEP users). The survey identified differences in intention among respondents based on social determinants such as education and migration status.(14) Research on previous MPXV outbreaks has shown that working with affected communities and leveraging digital media can support community uptake.(15-17) Although evidence on MPXV is only starting to emerge, structural barriers will remain a key issue in all outbreak responses.

This is the first UK co-produced community survey of this global public health emergency. It explores the understanding and acceptance of key public health messages about MPXV in the UK.

## MATERIALS AND METHODS

A cross-sectional anonymous survey was administered online between June 15 and July 27, 2022. The survey contained 25 questions exploring domains related to public health information: self-assessment of risk, acquisition of knowledge, understanding and acceptance of key public health information (origins of the virus, engagement with care, isolation rules and vaccine acceptability); views on media messaging with regards to MPXV in the UK. The survey was co-produced by the SHARE Collaborative at Queen Mary University of London (QMUL), and two community-based organisations involved in sexual health promotion and advocacy – The Love Tank CIC (mainly supports GBMSM, with a focus on men from racialised minorities, and migrant men) and the Sophia Forum (mainly supports women living with or at risk of HIV).

The anonymous survey was disseminated by The Love Tank CIC and Sophia Forum through their mailing lists and on social media channels (Facebook, Instagram, Twitter, and WhatsApp community groups) with further amplification by other community organisations representing communities at risk. The gay dating app Grindr provided a pro-bono broadcast link to the survey to its UK users on Friday 1 July 2022.

The survey was administered via a GDPR-compliant survey platform (SMART survey) and received ethical approval from the Queen Mary University Committee of Ethics panel (QMERC22.239, 6^th^ June 2022). The survey opening page contained information about the aims of the study and custodianship and use of study data. By clicking ‘continue’ and commencing the survey, individuals were considered to have given consent. Respondents could stop filling in the questionnaire at any time. Information regarding further support and information about MPXV was included.

Data analysis was undertaken by members of the QMUL research team (CM, JT, SP, CO, MS), with iterative feedback on results and review of all manuscript drafts from the remaining authors. The survey responses were analysed using Stata 17. There were varying levels of missing data by survey response, complete record analysis was implemented with the understanding that missing data may impact inferences (see Discussion). Basic descriptive statistics have been created for survey question responses and logistic regression has been implemented to explore associations. Odds ratio p values have been given for completeness but have not been used to statistically test hypotheses as no formal power calculation was implemented and due to the exploratory nature of the analyses, type I inflation would likely be present due to multiple testing. Models are presented as unadjusted, with inferences created to reflect this approach.

The study was not funded.

## RESULTS

1932 respondents completed the survey, 1691 (90%) were cis or trans men who have sex with men. In terms of gender identity, 1750/1932 identified as men, 88 as women and 64 as gender non-conforming (12 transgender men, 15 transgender women, 34 non-binary people). 1453 (77%) described their sexual identity as gay/ lesbian, 64 (3%) as queer, 221 as bisexual (12%), 81/1932 (4%) as heterosexual, 37 (2%) as pansexual. 744 (39%) were under the age of 40. 71% (1366) self-identified as being from White, 3% (67) as Black, 158 as Asian (8%), 2% as LatinX (36), 11% (203) as ‘Mixed or Other’ heritage groups. Employment was high at 85% and 79% had completed higher education. 137 (7%) identified themselves as living with HIV when asked if they lived with a disability or long-term condition. (Table 1).

**Table1:** Demographics of survey responses

| Demographic | Category | Frequency and percentage n(%). |
| --- | --- | --- |
| **MSM N=1886** | No | 195 (10.34) |
|  | Yes | 1691 (89.66) |
| **Age N=1911** | 18-24 | 114 (5.97) |
|  | 25-29 | 166 (8.69) |
|  | 30-39 | 464 (24.28) |
|  | 40-49 | 502 (26.27) |
|  | 50-59 | 445 (23.29) |
|  | 60+ | 220 (11.51) |
| **Gender Identify N=1905** | Cis Woman | 88 (4.62) |
|  | Cis Man | 1750 (91.86) |
|  | Transgender Woman | 15 (0.79) |
|  | Transgender Man | 12 (0.63) |
|  | Non-binary | 34 (1.78) |
|  | Other | 6 (0.31) |
| **Assigned at Birth N=1895** | Male | 1781 (93.98) |
|  | Female | 112 (5.91) |
|  | Intersex | 2 (0.11) |
| **Current Sexual Identity N=1890** | Straight (Heterosexual) | 81 (4.29) |
|  | Bisexual | 221 (11.69) |
|  | Gay/Lesbian | 1453 (76.88) |
|  | Pansexual | 37 (1.96) |
|  | Same gender loving | 14 (0.74) |
|  | Queer | 64 (3.39) |
|  | Other | 20 (1.06) |
| **Highest Level of Completed Education N=1859** | Completed school education | 304 (16.35) |
|  | Completed a Higher Education degree | 1472 (79.18) |
|  | No qualification | 27 (1.45) |
|  | Other | 56 (3.01) |
| **Current Work Situation N=1854** | Employed/ self-employed full time | 1385 (74.7) |
|  | Employed/ self-employed part time | 188 (10.14) |
|  | Student | 66 (3.56) |
|  | Not employed and on benefits | 50 (2.70) |
|  | Not employed and NOT entitled to benefits | 75 (4.05) |
|  | Other | 90 (4.85) |
| **Afford the Basic Needs for Yourself / your Household N=1840** | No | 166 (9.02) |
|  | Yes | 1674 (90.98) |
| **Consider Myself to Have a Disability N=1868** | No | 1571 (84.10) |
|  | Yes | 297 (15.90) |
|  | -Yes with HIV | 137 (7.09) |
| **Currently Living with Any Long-Term Illnesses N=1851** | No | 1284 (69.37) |
|  | Yes | 567 (30.63) |
| **Heritage group N=1932** | White | 1366 (70.70) |
|  | Black | 67 (3.47) |
|  | Asian | 158 (8.18) |
|  | Latinx | 37 (1.92) |
|  | Other, Not Disclosed | 203 (10.51) |

Up to and during the survey period (June and July 2022), 49.5% (955) had heard/read a lot about MPXV, 48.8% (941) had heard/read little and 1.6% (31) did not know about it. The top three places that people reported hearing about MPXV were online news 1099 (57%), TV or radio news (51%) and printed press (27%). Amongst the social media platforms, Twitter was the most common source of MPXV information (21%), followed by Facebook (12%), Instagram (5%) and snapchat (0.4%). Other reported sources of information were health care professionals (HCPs) (11%), dating apps (13%) and sex on premises venues (2%).

HCPs were the most trusted source of information, ranking highest among 713 (36%) respondents, followed by government websites 552 (29%), Google searches 183 (9%) and community organisations 82 (4%). Within the top two most trusted sources, trust varied widely with respect to sexuality (MSM vs non MSM), gender, gender identity, age and education. (Table 2).

**Table 2:** Frequency and percentage reporting trust in healthcare providers and government websites.

|  |  | I Trust Healthcare Providers<br>n(%) | I Trust Government Websites<br>n(%) |
| --- | --- | --- | --- |
| **MSM** | No | 72 (36.92) | 34 (17.44) |
|  | Yes | 627 (37.08) | 512 (30.28) |
| **Age** | >=40 | 415 (35.56) | 319 (27.34) |
|  | <40 | 291 (39.11) | 231 (31.05) |
| **Identify as** | Cis- Man, Transgender Man | 1114 (36.78) | 518 (29.40) |
|  | Cis- Woman, Transgender Woman | 42 (40.78) | 20 (19.42) |
|  | Non-Binary | 12 (35.29) | 10 (29.41) |
|  | Other | 3 (50.00) | 1 (16.67) |
| **I was assigned at birth:** | Male | 651 (36.55) | 532 (29.87) |
|  | Female | 47 (41.96) | 18 (16.07) |
| **Sexual Identity** | Gay/Lesbian, Queer, Same gender loving | 944 (38.34) | 455 (29.72) |
|  | Bisexual | 65 (29.41) | 64 (28.96) |
|  | Straight (Heterosexual) | 28 (34.57) | 17 (20.99) |
|  | Other | 23 (40.35) | 10 (17.54) |
| **Highest education level** | Completed a Higher Education degree | 575 (37.93) | 452 (29.82) |
|  | Other | 112 (32.65) | 89 (25.95) |
| **Current Work situation** | Employed | 584 (37.10) | 465 (29.54) |
|  | Other | 97 (34.64) | 80 (28.57) |
| **Afford Basics** | Yes | 629 (37.57) | 496 (29.63) |
|  | No | 53 (31.93) | 38 (22.89) |
| **Disability** | No | 589 (37.49) | 469 (29.85) |
|  | Yes | 103 (34.68) | 70 (23.57) |
| **Long Term Illness** | No | 482 (37.54) | 392 (30.53) |
|  | Yes | 205 (36.16) | 146 (25.75) |
| **Heritage group** | White | 548 (40.12) | 373 (27.31) |
|  | Black | 21 (31.34) | 20 (29.85) |
|  | Asian | 44 (27.85) | 59 (37.34) |
|  | Mixed | 64 (31.53) | 56 (27.59) |
|  | Latinx | 11 (29.73) | 16 (43.24) |
|  | Other, No Disclosed | 25 (24.75) | 28 (27.72) |

Table 2 shows that people assigned female at birth have a lot more trust in health care providers (42%) compared to people assigned male (37%). However, this is reversed for government websites with 30% people assigned male at birth trusting them compared to only 16% for female assigned at birth. There were also differences regarding trust in HCPs based on heritage with White heritage groups being most trusting of HCPs.

34% reported not understanding the current public health information very well, around half understood it ‘fairly well’ and only 17% reported understanding it ‘very well’ (Table 3). More men (cis and trans men) vs women (cis and trans women) said that they understood the information ‘very well’ (17% vs 8%). More LatinX people said they understood the information ‘very well’ than people from Black heritage groups (35% vs 12%). People who said they could not afford basic needs and those without a university degree were less likely to say that they had understood the information ‘very well’.

**Table 3:** Reponses to how well do you feel you understand current public health information about monkeypox?

|  |  | Not very well | Fairly well | Very well |
| --- | --- | --- | --- | --- |
| **MSM** | No | 63 (32.98) | 100 (52.36) | 28 (14.66) |
|  | Yes | 574 (34.35) | 811 (48.53) | 286 (17.12) |
| **Age** | >=40 | 362 (31.4) | 575 (49.87) | 216 (18.73) |
|  | <40 | 281 (38.39) | 348 (47.54) | 103 (14.07) |
| **Identify as** | Cis- Man, Transgender Man | 596 (34.23) | 843 (48.42) | 302 (17.35) |
|  | Cis- Woman, Transgender Woman | 36 (36.00) | 56 (56.00) | 8 (8.00) |
|  | Non-Binary | 7 (21.21) | 19 (57.58) | 7 (21.21) |
|  | Other | 2 (33.33) | 4 (66.67) | 0 (0.00) |
| **I was assigned at birth:** | Male | 601 (34.17) | 854 (48.55) | 304 (17.28) |
|  | Female | 38 (34.55) | 60 (54.55) | 12 (10.91) |
|  | Intersex | 0 (0.00) | 1 (50.00) | 1 (50.00) |
| **Sexual Identity** | Gay/Lesbian, Queer, Same gender loving | 513 (33.93) | 744 (49.21) | 255 (16.87) |
|  | Bisexual | 82 (37.44) | 98 (44.75) | 39 (17.81) |
|  | Straight (Heterosexual) | 27 (33.75) | 43 (53.75) | 10 (12.50) |
|  | Other | 638 (34.19) | 912 (48.87) | 316 (16.93) |
| **Highest education level** | Completed a Higher Education degree | 487 (32.45) | 751 (50.03) | 263 (17.52) |
|  | Other | 135 (40.18) | 148 (44.05) | 53 (15.77) |
| **Current Work situation** | Employed | 522 (33.55) | 762 (48.97) | 272 (17.48) |
|  | Other | 102 (37.23) | 133 (48.54) | 39 (14.23) |
| **Afford Basics** | Yes | 536 (32.35) | 827 (49.91) | 294 (17.74) |
|  | No | 79 (48.77) | 64 (39.51) | 19 (11.73) |
| **Disability** | No | 523 (33.74) | 767 (49.48) | 260 (16.77) |
|  | Yes | 107 (36.52) | 131 (44.71) | 55 (18.77) |
| **Long Term Illness** | No | 425 (33.46) | 623 (49.06) | 222 (17.48) |
|  | Yes | 200 (35.91) | 265 (47.58) | 92 (16.52) |
| **Heritage group** | White | 438 (32.54) | 681 (50.59) | 227 (16.86) |
|  | Black | 25 (37.88) | 33 (50.00) | 8 (12.12) |
|  | Asian | 60 (38.71) | 75 (48.39) | 20 (12.90) |
|  | Mixed | 81 (40.10) | 86 (42.57) | 35 (17.33) |
|  | Latinx | 10 (27.03) | 14 (37.84) | 13 (35.14) |
|  | Other, No Disclosed | 34 (35.79) | 42 (44.21) | 19 (20.00) |

21% did not know if they were at risk, around half (51%) considered themselves at risk and 27% did not consider themselves at risk. 982 (66%) of respondents reported some level of personal risk concern about MPXV, and 70% of MSM were more concerned about getting MPXV compared to only 37% in non-MSM (odds ratio 1.38 (95%CI 1.02 – 1.74).

60% (1147/1911) ‘agreed’ or ‘somewhat agreed’ that MPXV originates from animals, 31% (597) neither agreed nor disagreed, 114 (6%) disagreed with this statement. People who could not afford basic needs compared to those than could [OR: 0.48 95%CI (0.35 - 0.67) and those without a higher education degree compared to responders with a degree [OR: 0.74 95%CI (0.58 - 0.94)] were less likely to ‘agree’ or ‘somewhat agree’ that MPXV came from animals. MSM respondents were far more likely to say they ‘agree’ or ‘somewhat agree’ that MPXV came from animals than non-MSM OR:1.52 95%CI (1.13 - 2.05). Table 4 shows the odds ratios for each demographic group unadjusted.

**Table 4:**
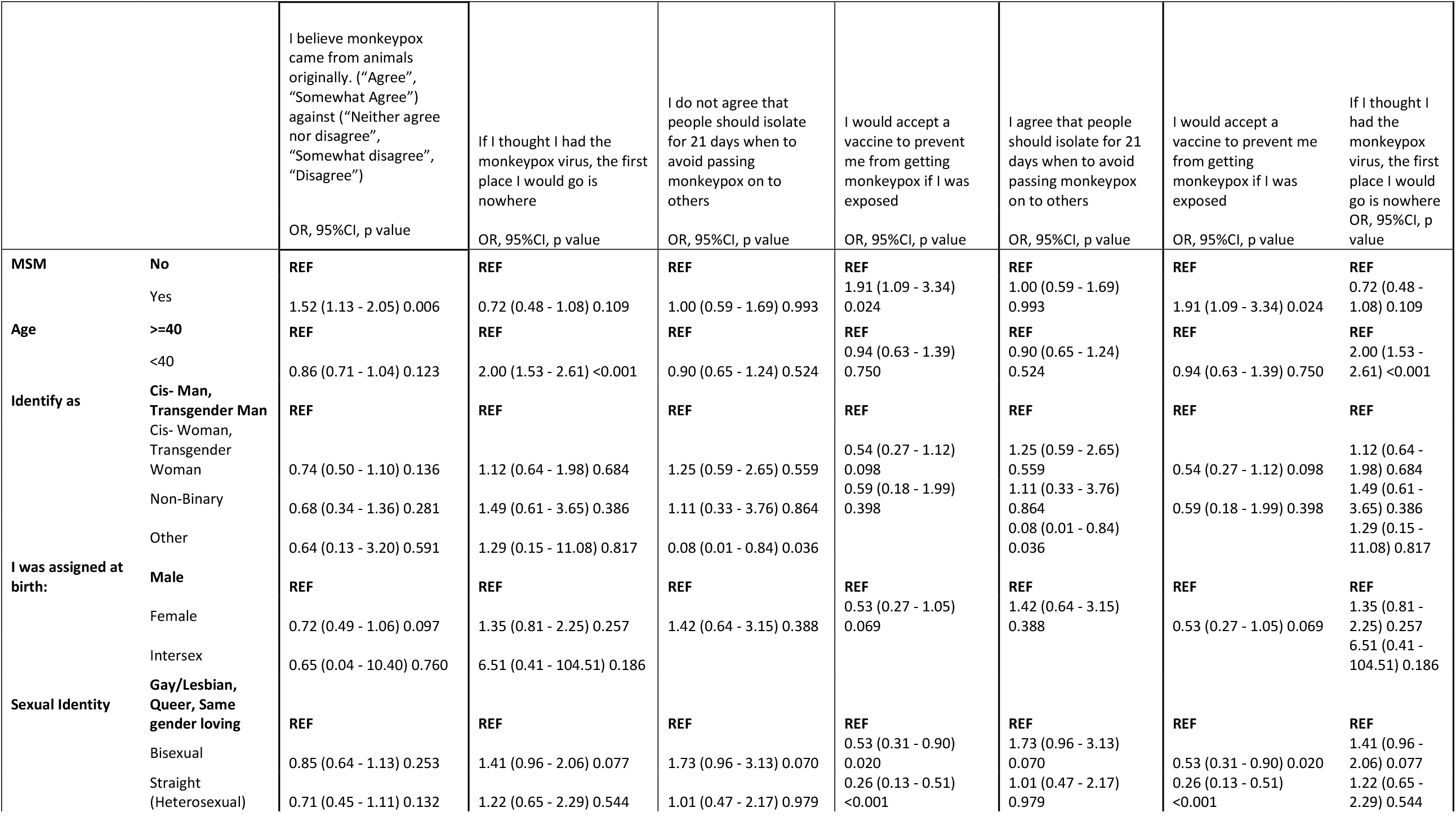
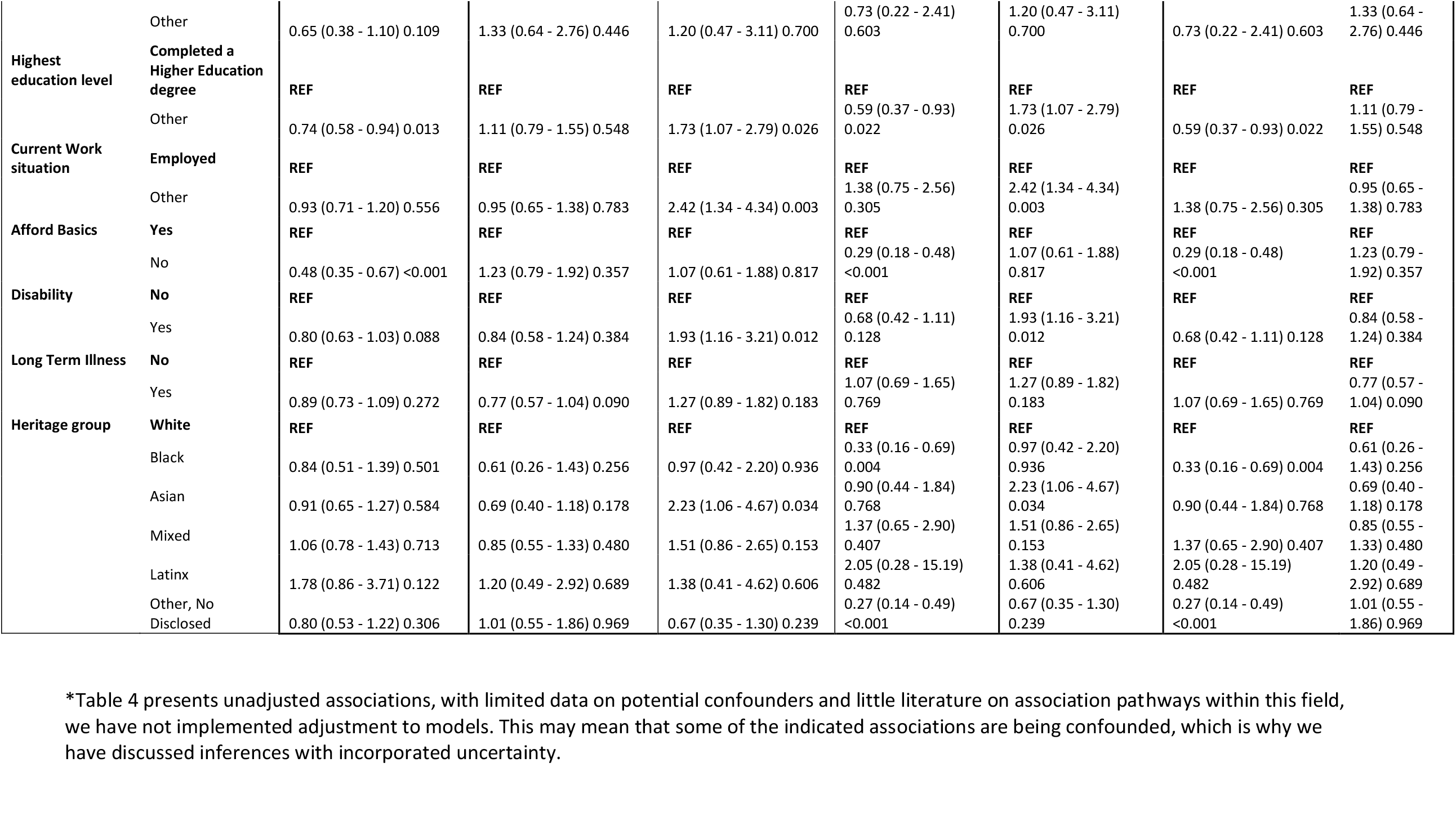
Unadjusted logistic regressions exploring the association between demographics and the four proxy questions to assess the efficacy of the public health messaging*.

Half (951) responded that they would attend a sexual health clinic if they thought they had MPXV, the remainder said they would attend their GP 554 (29%), accident and emergency department 107(6%), a pharmacy 23 (1%), and 9 (0.5%) a voluntary organisation. 260 (14%) replied that they would go ‘nowhere’. Notably, those under the age of 40 were most likely to say they would go ‘nowhere’ (ie not to engage with healthcare) if they had MPXV compared to respondents aged over 40 years [OR: 2.00 95%CI (1.53 - 2.61), see Table 4. There was little difference in responses between the MSM and non-MSM respondents.

1184 (62%) agreed that people should isolate for 21 days to avoid passing on MPXV to others, 181 (9%) disagreed, and 556(29%) were not sure. Factors associated with not agreeing that people should isolate appeared to be connected to not having a higher education degree compare to respondents that did OR: 1.73 95%CI(1.07 - 2.79), not being employed compare to those that are OR: 2.42 95%CI(1.34 - 4.34), having a disability compared to not OR:1.93 95%(1.16 - 3.21) and identifying as being from Asian compared to White heritage groups OR: 2.23 (1.06 - 4.67). There appeared to be little difference between MSM and non-MSM respondents, see table 4.

1659 (86%) reported that they would accept a vaccine for MPXV if offered, 114 (6%) would not, 149 (8%) were not sure. MSM respondents were more likely to answer that they would accept a vaccine compared with non-MSM OR: 1.91 95%CI(1.09 - 3.34). Factors that appeared to be associated with a lower intention to take up a vaccine include identifying as bisexual [OR: 0.53 95%CI(0.31 - 0.90)], or heterosexual [OR: 0.26 95%CI(0.13 - 0.51)] compared to gay/lesbian, queer or same gender loving. Similarly, those that could not afford basic needs expressed lower intention to take the vaccine OR: 0.29 95%CI(0.18 - 0.48). Compared to respondents that identify as being from White heritage groups, respondents identifying as being from Black and ‘other/ did not disclose’ heritage groups, also appeared less inclined to take up the vaccine (OR:0.33 95%CI(0.16 - 0.69) OR:0.27, 95%CI(0.14 - 0.49) respectively). There was little evidence of response difference between other demographic groups (results in Table 4).

45% of respondents, regardless of sexual identity status, said that they did not feel people who talk about MPXV in the media represent them. There was little difference in response between MSM and non-MSM respondents, suggesting that people generally did not feel well represented in the media response.

## DISCUSSION

This was the first UK co-produced community survey assessing public views on the early phase of the public health response to the MPXV outbreak. It reflects predominantly the perspectives of GBMSM - the most affected community in this outbreak. Respondents were highly educated and reported high levels of understanding and awareness of MPXV. Key findings point to shortcomings in the public health responses in the early phase of the MPXV outbreak in the UK, especially among groups who already face social and structural barriers to care.

Sources of trusted information were not uniform within the surveyed population: women were more trusting of HCPs and less trusting of government websites. Those from White heritage groups were more likely to trust government websites than people of other ethnicities.

With respect to understanding of the public health information on MPXV, differences based on social determinants, such as lack of higher education and unemployment, were associated with worse self-assessed understanding of public health information.

The concerning number (40%) of respondents that did not answer that they believed in the zoonotic origins of MPXV suggests confusion, lack of information, and the potential circulation of conspiracy theories about the causes of the MPXV outbreak. This finding concurs with the findings from the recent large US survey of the general population in which concerning levels of misinformation and beliefs in conspiracy theories were identified.(12)

Key UK public health messages covered where to attend if an individual developed MPXV symptoms. This is a particularly important piece of information for the public health response to infectious disease emergencies. Members of the public have been advised to attend sexual health clinics if symptomatic with MPXV, according to the websites of the Department of Health and Social Care, NHS, and UKHSA. However, only 49.5% survey respondents understood or accepted this advice. There is a need to improve awareness of and inclusive access to appropriate help-seeking during outbreaks of infectious diseases. A&E and primary care clinics continue to be seen as first-choice options, which could introduce challenges for the ongoing response, given the infection control issues that attending these clinics would entail.

Differences in intentions to attend clinics were observed across respondents from different heritage groups, which may reflect broader health inequalities in the UK as observed amongst racially minoritised communities with regards to relevant services including sexual health and immunisation. Working with multiple communities to explore access to other spaces to receive care, given current health inequalities and the burden on sexual health services, helps to address structural barriers that continue to be experienced for a variety of reasons. (An example of this were successful outreach vaccination events offered at places like Black Pride in London in August 2022).

The majority of respondents agreed that people should isolate for 21 days if they acquire MPXV. However, social determinants, such as ability, education, and unemployment once again negatively affected people’s intentions around willingness to self-isolate. As we witnessed during COVID-19 response in high-income liberal democracies, conditions of employment and housing elevate risk and limit preventive options, restricting the ability to avoid close contact and isolate after infection.(18) It is likely that isolation-related experiences during the recent COVID-19 pandemic had an impact on these findings.

The study shows very high vaccine acceptability amongst both those who did and those who did not perceive themselves to be at risk for infection. This is a striking finding as the survey was conducted at a time where vaccines were not yet widely available and very little information was in circulation about them including about how they work, where they could be accessed and by whom. However, given the demographic characteristics of the sample, there is a need to consider the potential for health inequalities amongst the minority of respondents unwilling to be vaccinated; in this study respondents from Black heritage groups were less likely to accept vaccines than their White counterparts. People who could not afford basic needs were also less likely to take up vaccines.

It is plausible the attitudes towards MPXV immunisation will in time reveal similarities to what was witnessed in relation to COVID-19 vaccines. Certainly, legacies of racism and prejudice also colour vaccine awareness and acceptability. (1, 19) Community-based health promotion and a diversity of interventions (not only clinic-centred) are central to an inclusive immunisation response if it is to reach those most affected.(20)

The media coverage was not considered representative of the most affected groups. Given that most people reported hearing about MPXV via traditional media this is of significant concern and can be improved by collaboration and co-production with different communities to shape communication strategies.

### Strengths and weaknesses of the survey analysis

To our knowledge, this is the first community co-produced cross-sectional survey of awareness, understanding, and response among affected communities during the 2022 MPXV emergency. Like the Annenberg survey of the general population in the US and Dutch MSM survey, we found misunderstandings of key public health information.

The study sample is overwhelmingly White, male, educated and in employment, and while it is broadly reflective of the population affected by MPXV in the UK, it is not a representative sample against the whole population. Survey data also contained missing data, which is unlikely to be missing at random and thus we are limited to conclude only on the data survey respondents wished to give. However, the study had the advantage of being entirely co-produced and distributed through key community organisations who are at the forefront of the MPXV response in the UK. This meant that the study was able to reach a large sample of potentially highly affected community members in a short period of time.

Moreover, the sample included a significant proportion of women and gender non-conforming respondents in comparison to similar surveys usually recruiting MSM, as well as a diversity of ethnic heritage groups which allowed for some comparisons. The survey was designed to respond rapidly to the need to understand the evolving situation in June and July and the large recruitment is evidence of the high levels of interest and concerns amongst affected communities at the time.

## Conclusions

There is a need to maintain vigilance about media representations during outbreak and ongoing response. Public health officials are not responsible for the reaction of media outlets to new disease outbreaks. However, there is a need to carefully consider the management of information given through the media and who is selected to represent the public health response. A very low number of respondents felt represented in any way by people they saw in the media discussing MPXV. This is likely to exacerbate mistrust and foster further misunderstanding. It is essential to include a diversity of members of different affected communities in media interventions.

It is of the highest importance to ensure information and resources for both public health and media dissemination are co-produced with most at-risk communities, including racially minoritised, gender non-conforming and younger MSM groups. It is also very important to consider the effects of social mobility, poverty and wider social and structural determinants of health and their immediate effect on healthcare engagement and access.

Further research is needed to establish awareness, risk perception, engagement in care, and vaccine acceptability, especially in more diverse and vulnerable populations. Lessons from previous viral outbreaks and pandemics evidence the need to respond to in ways that address rather than exacerbate health inequalities during new infectious disease emergencies.

## Data Availability

All data produced in the present study are available upon reasonable request to the authors

## References

1. Willis DE, Andersen JA, Montgomery BEE, Selig JP, Shah SK, Zaller N, et al. COVID-19 Vaccine Hesitancy and Experiences of Discrimination Among Black Adults. J Racial Ethn Health Disparities. 2022:1–10.

2. Bogart LM, Wagner GJ, Galvan FH, Landrine H, Klein DJ, Sticklor LA. Perceived Discrimination and Mental Health Symptoms among Black Men with HIV. Cultur Divers Ethnic Minor Psychol. 2011;17(3):295–302.

3. English D, Carter JA, Forbes N, Bowleg L, Malebranche D, Talan A, et al. Intersectional discrimination, positive feelings, and health indicators among Black sexual minority men. Health Psychol. 2020;39(3):220–9.

4. Sotgiu G, Dobler CC. Social stigma in the time of coronavirus disease 2019. European Respiratory Journal. 2020;56(2).

5. Whitacre R, Oni-Orisan A, Gaber N, Martinez C, Buchbinder L, Herd D, et al. COVID-19 and the political geography of racialisation: Ethnographic cases in San Francisco, Los Angeles and Detroit. Global Public Health. 2021;0(0):1–15.

6. Baldassarre A, Giorgi G, Alessio F, Lulli LG, Arcangeli G, Mucci N. Stigma and Discrimination (SAD) at the Time of the SARS-CoV-2 Pandemic. Int J Environ Res Public Health. 2020;17(17):6341.

7. Daskalakis D, Paul M, Leandro M, Jonathan M. Monkeypox: Avoiding the Mistakes of Past Infectious Disease Epidemics | Annals of Internal Medicine. 2022.

8. Ayala G, Sprague L, Merwe LL-Avd, Thomas RM, Chang J, Arreola S, et al. Peer- and community-led responses to HIV: A scoping review. PLOS ONE. 2021;16(12):e0260555.

9. Hansen H, Metzl J. Structural Competency in the U.S. Healthcare Crisis: Putting Social and Policy Interventions Into Clinical Practice. J Bioeth Inq. 2016;13(2):179–83.

10. Metzl JM, Hansen H. Structural competency: Theorizing a new medical engagement with stigma and inequality. Social Science & Medicine. 2014;103:126–33.

11. Harvey M, Neff J, Knight KR, Mukherjee JS, Shamasunder S, Le PV, et al. Structural competency and global health education. Global Public Health. 2020:1–22.

12. Annenberg Public Policy C. The Annenberg Public Policy Center of the University of Pennsylvania [Internet]2022 2022/07/29/T12:12:42+00:00. Available from: https://www.annenbergpublicpolicycenter.org/survey-1-in-5-americans-fear-getting-monkeypox-but-many-know-little-about-it/.

13. Ortiz-Martínez Y, Galvis-Cataño LM, Arias-Rodríguez D, Romero-Dager C, Bonilla-Aldana DK, Rodriguez-Morales AJ. YouTube and 2022 Monkeypox outbreak: opportunities for awareness and infection control. Journal of Hospital Infection. 2022;0(0).

14. Wang H, Paulo KJIdAd, Gültzow T, Zimmermann HML, Jonas KJ. Monkeypox self-diagnosis abilities, determinants of vaccination and self-isolation intention after diagnosis among MSM, the Netherlands, July 2022. Eurosurveillance. 2022;27(33):2200603.

15. Reed KD. Monkeypox, Marshfield Clinic and the Internet: Leveraging Information Technology for Public Health. Clin Med Res. 2004;2(1):1–3.

16. Roess AA, Monroe BP, Kinzoni EA, Gallagher S, Ibata SR, Badinga N, et al. Assessing the Effectiveness of a Community Intervention for Monkeypox Prevention in the Congo Basin. PLoS Negl Trop Dis. 2011;5(10):e1356.

17. Ogoina D, Izibewule JH, Ogunleye A, Ederiane E, Anebonam U, Neni A, et al. The 2017 human monkeypox outbreak in Nigeria—Report of outbreak experience and response in the Niger Delta University Teaching Hospital, Bayelsa State, Nigeria. PLOS ONE. 2019;14(4):e0214229.

18. Hengel KMO, Burdorf A, Pronk A, Schlünssen V, Stokholm ZA, Kolstad HA, et al. Exposure to a SARS-CoV-2 infection at work: development of an international job exposure matrix (COVID-19-JEM). Scand J Work Environ Health. 2022;48(1):61–70.

19. Kricorian K, Turner K. COVID-19 Vaccine Acceptance and Beliefs among Black and Hispanic Americans. PLOS ONE. 2021;16(8):e0256122.

20. Daskalakis D. Monkeypox: Outbreak and response in non-endemic countries. AIDS 2022, Montreal, Quebec, CA.

